# No evidence of progressive pro-inflammatory cytokine storm in brain dead organ donors - A time course analysis using clinical samples

**DOI:** 10.1101/2022.02.16.22270687

**Authors:** Kasia D Bera, Joel Tabak, Rutger J Ploeg

## Abstract

Organ donation after brain death (DBD) is an important source of transplanted organs as chronic undersupply means many patients die whilst awaiting a transplant. Improving organ quality and graft survival is key to reducing waiting lists. The interval between brain death confirmation and organ procurement offers an opportunity to reduce organ injury or encourage repair, yet data is lacking on the effects of brain injury on future grafts and which pathways to target. In fact, the consensus view has been that brain death exposes organs to a progressively pro-inflammatory environment, suggesting organ procurement must occur within a limited time window. Here, we developed a novel method of studying time-course changes in serum proteins in a DBD cohort from a UK biobank. Individual donor trajectories were combined to determine the time course of pro-inflammatory (Tumour Necrosis Factor alpha, Interleukin 6, Complement) mediators and markers of central nervous injury (Neuron Specific Enolase, Glial Fibrillary Acidic Protein) after confirmation of brain death. We found no evidence of a rise of TNF-alpha, NSE, IL-6 or C5a with prolonged duration of brain death, questioning the consensus view of progressively pro-inflammatory environment. We also identified complement as a potential target for therapeutic intervention to improve organ quality.

## 1. Introduction

Organ transplantation is a lifesaving and cost-effective treatment for patients with end-stage organ failure; however, donor organ shortages mean many patients still die whilst awaiting a transplant. In addition, many procured older and higher risk donor organs are considered un-transplantable, declined by transplant centres and thus not utilised. It is therefore imperative that we improve organ quality as well as maintaining good long-term transplant survival. Donation after brain death (DBD) is the most common source of deceased donor organs worldwide. Whilst DBD offers a more controlled environment than procuring organs after circulatory death (when donors will have suffered from a respiratory and cardiac arrest) long term outcomes between both types of donation remain comparable^1,2^. Historically, the events surrounding brain death have been described as ‘hostile’ including a catecholamine storm with significant haemodynamic, metabolic and hormonal changes as well as a progressive release of pro-inflammatory mediators (e.g Tumour necrosis Factor (TNF) -alpha and Interleukin (IL)-6) and activation of the complement cascade. Chemokine exposure contributes to the long-term trajectory of an organ, leading to fibrosis, impacting on long-term function and ultimately reducing graft survival^3–5^.

To minimise the duration of grafts-to-be in a presumed hostile environment, rapid procurement of organs was adopted. Retrospective analyses, however, demonstrate that longer duration of organ donor management in critical care actually may be beneficial for some of the transplanted organs, reducing rate of delayed graft function (DGF) for renal allografts from younger donors and without negative impact on transplanted liver or pancreas^6,7^. The period of brain dead organ donor management in critical care also offers a therapeutic window of intervention, yet our understanding of inflammatory processes during this period remains limited. Management of the brain dead donor has evolved and improved in recent history: for example, in the UK a ‘donor care bundle’ used by intensivists provides guidance and donors universally receive corticosteroids^8^, yet detailed knowledge of how this impacts the pro- and anti-inflammatory balance is currently lacking. Animal models of brain death have been limited to a short time frame (4-6 hours)^9–12^. Prior studies of inflammatory serum changes in DBD organ donors measured only a limited number of time points and/or study donors with heterogenous pathologies that result in brain death^13–18^. Understanding details of the time course surrounding brain death is key to developing strategies to reduce organ injury or promote repair, but detailed translation from preclinical studies is lacking and availability of clinical samples is limited.

We hypothesised that the selection of a donor cohort with a common underlying pathology would enable us to determine the temporal changes of biomarkers of brain injury (NSE, GFAP – to reflect neuronal and glial injury, respectively) and pro-inflammatory mediators (TNF-a, IL-6 and complement), from confirmation of brain death through to organ procurement. Understanding the time course can be then used to identify target pathways for treatment even before organ procurement to improve long term organ quality and graft survival.

## 2. Methods

### 2.1 Donor cohort and serum sample selection process

Serum samples from the UK Quality in Organ Donation (QUOD) biobank were selected to study the brain death period between 10 and 30 hours (from confirmation of brain death by completed second brain stem test through to organ procurement). Inclusion criteria used were: presence of valid consent; availability of three serum samples (before brain death; after confirmation of brain death, at end of donor management period); intracranial haemorrhage (ICH) as cause leading to brain death, as well as the documented duration of brain death. The following exclusion criteria were used: incomplete set of serum samples; other causes of brain death (such as but not limited to hypoxic brain injury, trauma, ischaemia, infection); documented systemic source of elevated inflammatory markers (trauma, documented infection such as e.g. pneumonia or urinary infection).

To allow selection of donors who were similar in characteristics other than the duration of brain death, the 20-hour period was broken down into four 5-hour blocks (10-15 hours, 15-20 hours, 20-25 hours, 25-30 hours of brain death) and for each block, donors were selected to be balanced for age, gender, body mass index, co-morbidities (especially hypertension and diabetes), duration of cold ischemia time and renal function after transplantation. The inclusion of the latter two ensured that the selection of donors was not biased to those with known factors associated with particularly good or bad post-transplant outcomes. In addition, donors who fulfilled all inclusion criteria but had documented ‘extremes’ of brain death (< 10h or > 30h) were also included to allow study of time course either side of the core time frame.

The final cohort included n=27 donors. All included donors had a complete set of serum samples, however for six donors the exact time point of the first sample collection at time of admission was not documented; the time and date of admission recorded by NHSBT was used instead. These donors with one missing time point were evenly distributed between the brain death groups. The QUOD programme research approval as a Research Tissue Bank (REC ref 13/NW/0017, from North West - Greater Manchester Central Research Ethics Committee,) covers the provision of data and research samples for research into improving the quality of organ quality for transplantation.

### 2.2 Enzyme linked immune absorption assay (ELISA)

All received samples were stored at -80C before use. DuoSet® ELISA Kits (R&D Systems) for human IL-6, TNF-alpha, Complement 5a, NSE and GFAP were used according to the manufacturer’s instructions. Each sample was measured in duplicates using a Bio-Rad iMark™ Microplate Reader. MatLab and GraphPad Prism9 were used for data analysis. The mean value from the duplicate measurements (followed by transformation using ln (Y+1)) was used for the time course plots (Fig 1).

**Figure 1.**
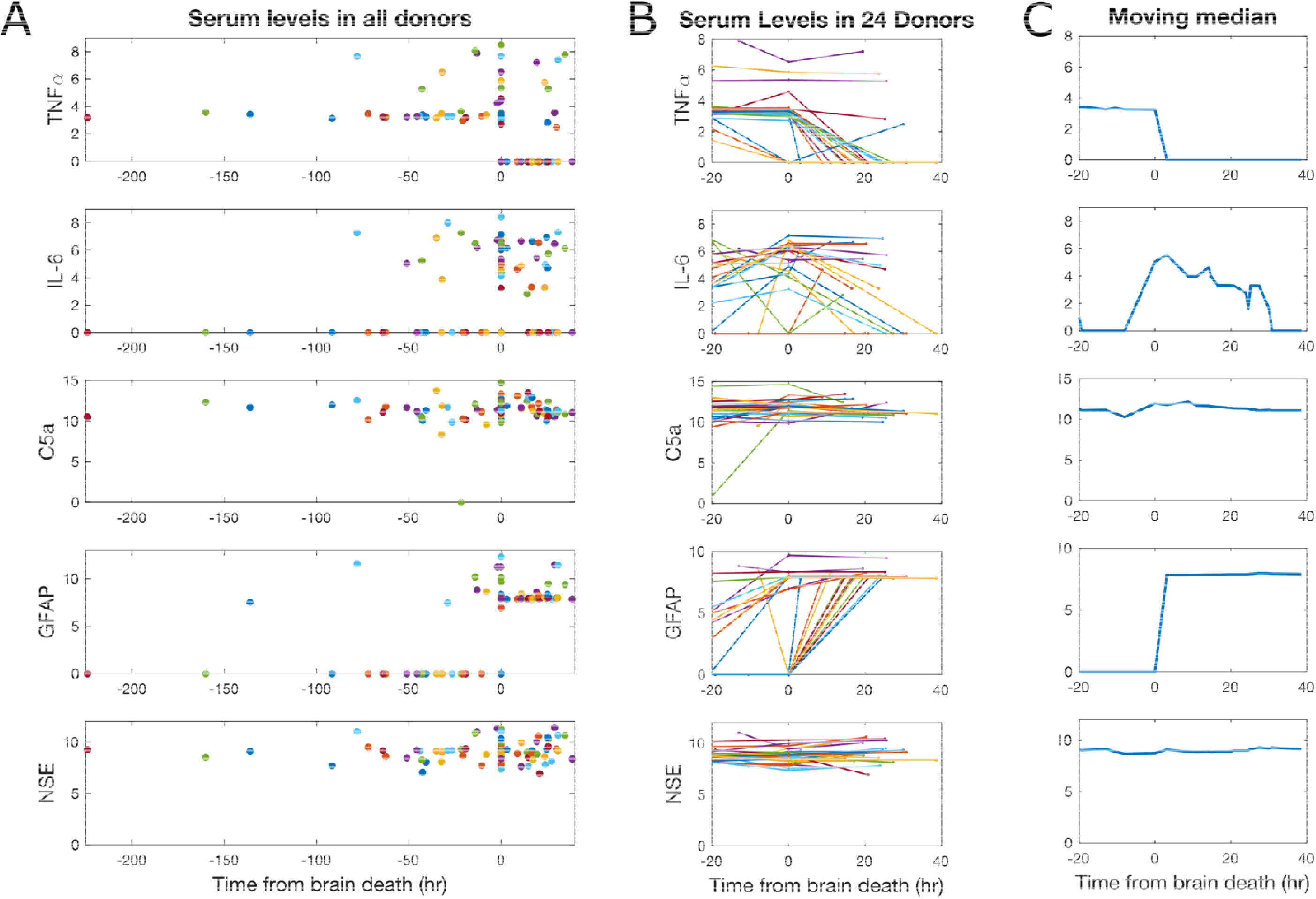
Serum time course analysis. Each row represents serum levels for one of the studied biomarkers: Tumour Necrosis Factor alpha (TNF⍰, Interleukin-6 (IL-6), Complement C5a, Glial Fibrillary Acidic Protein (GFAP) and Neuronal Specific Enolase (NSE), all levels shown in ln(y+1). For each donor, three samples based on clinically defined time points (DB1, DB2, D3) were measured, DB2 was set as t=0 to allow comparison. Panels A, B, C show analysis of serum levels from individual sample levels to combined time course analysis. (A) Comparison of serum levels of the whole donor cohort, each donor in different colour. (B) donor trajectories of final donor cohort (connecting lines depict individual donor trajectories). (C) calculated moving median to demonstrate time course for each biomarker (moving median window of 20 hours).

### 2.3 Moving median analysis

Before combined analysis of donor serum levels, the Robust regression and Outliner removal (ROUT) method was used to identify outliers. Donors for which at least four outliers out of the five measured serum molecules were excluded before a moving median was calculated. This analysis removed three donors – one from the 25-30h brain death duration group and two from the >30h brain death group, representing the oldest donors in this group, one with a missing admission time point.

Subsequently, data for all remaining donors was combined using moving median plots for each serum level of the molecules. In detail, the time of brain death confirmation was defined as t = 0 for each donor. Thus, for each patient and each marker, we have a set of three sample measurements, one before brain death, one at brain death (t = 0), and one after brain death. Because the sample before and after brain death were obtained at different times for the different patients, the ensemble of measurements across patients contain information about marker time course before and after brain death. We did not directly combine all the measurements into one time course for each marker, as this would make the strong assumption that markers from different patients follow an identical time course. Here, we make a softer assumption that the time courses from different patients follow qualitatively *similar* trajectories. Following that assumption, we can combine measurements from different patients using a moving median of the time points, as long as the window is large enough to account for the variability across patients.

We constructed a moving median for the 20-hour period before confirmation of brain death, and the 40-hour period following brain death. For the 20-hour period before confirmation of brain death, we replaced each level by the median of all levels within a 20-hour window centred on that point and sampled before t = 0. Similarly, for the 40-hour period of time after confirmation brain death, we used a 20-hour moving window using only levels sampled after t = 0. For t = 0, the median of all samples collected at this clinical time point was used.

## 3. Results

### 3.1 Selection of study cohort: DBD donors with different durations of brain death

The Quality in Organ Donation (QUOD) biobank contains samples from over 85% of all deceased organ donors in the UK, alongside detailed clinical donor and recipient information^19^. Samples, including serum and urine, are collected at clinically pre-defined time points: admission (DB1), confirmation of brain death (DB2), end of donor management (DB3), and immediately prior to removal of the donor organs (DB4). As most DBD organ donors in the UK have suffered a non-survivable ICH, the selection of donors was limited to this pathology. Our selection criteria identified donors with varying duration of brain death, as defined by period from confirmation of brain death to end of organ donor management but before organ procurement. This created a cohort that included durations of brain death ranging from 10 to 30 hours. At the point of study design, the QUOD biobank included 268 DBD with a complete set of three serum samples (DB1-3); of those, 51 donors had ICH as the documented cause of brain death, and 45/51 donors (88.23%) had a documented brain death duration between 10 and 30 hours (mean 19.3 +/- 7 hours). We subsequently excluded donors with a history of other sources of inflammation. To further define the final study cohort, 5-hour windows of brain death duration were defined with five donors per group; within each group, donors were selected to be matched for factors such as body mass index, sex, and age. Two additional groups of donors with ‘extremes’ of duration of brain death were included: ‘short BD’ (<10 hours) with 2 donors and ‘long BD’ (>30 hours) with 5 donors. This resulted in n=27 selected donors for the final cohort. Characteristics of all included donors are displayed in Table 1. One-way ANOVA was used to evaluate the balance between the groups regarding the specified continuous parameters and confirm matched creation of study cohort. When the two extremes of BD groups are included, donor age was statistically different between all groups, with notable anti-correlation between donor age and brain death duration (p = 0.0017, R^2^ = 0.33). This likely reflects underlying decision making: e.g., for younger donors a prolonged duration of BD might be deemed acceptable, whilst organs from older donors might be accepted when cold ischaemia time and transport duration can be minimised.

**Table 1.**
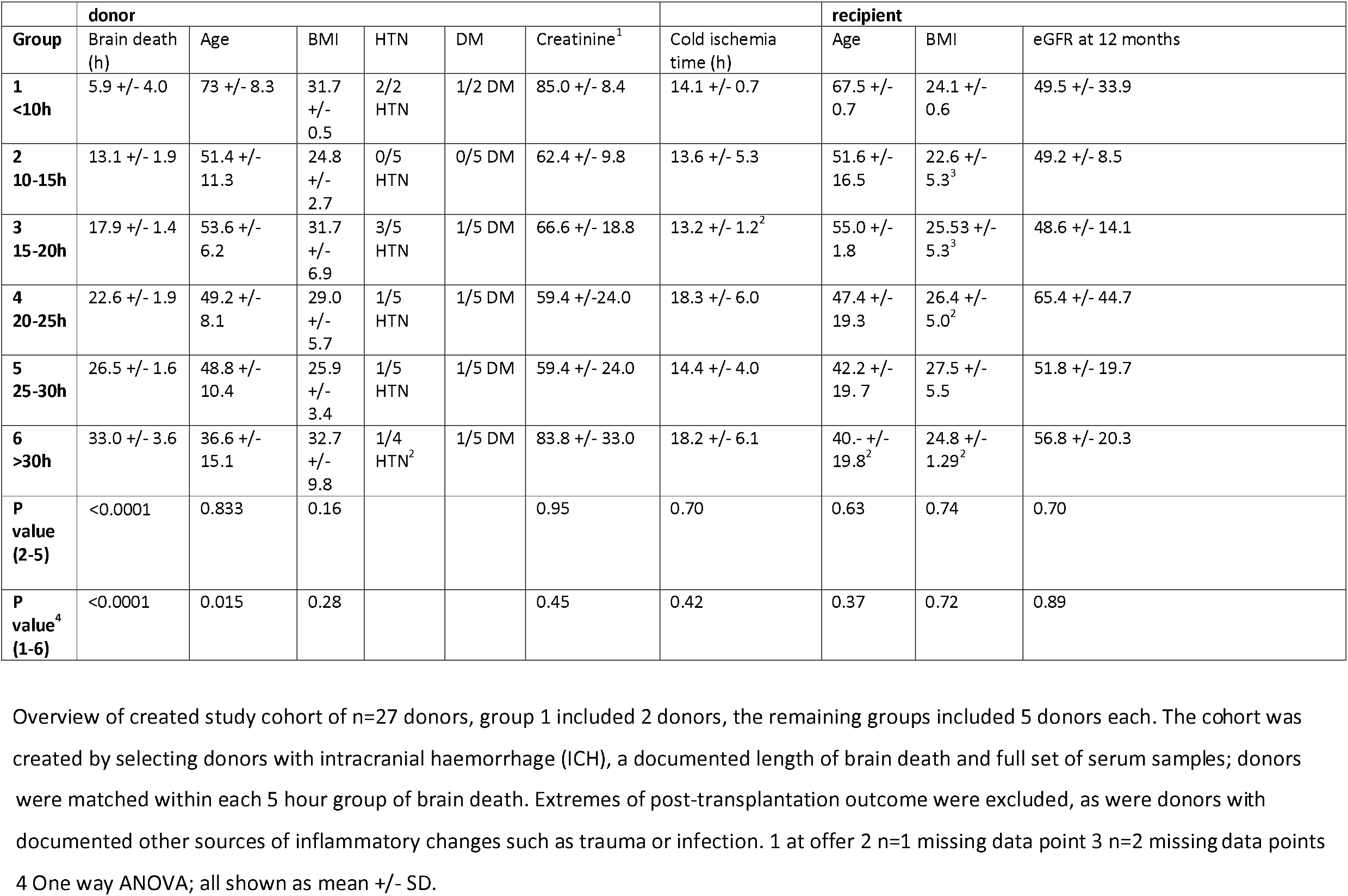
Clinical characteristics of final study cohort.

|  | donor |  |  |  |  |  |  | recipient |  |  |
| --- | --- | --- | --- | --- | --- | --- | --- | --- | --- | --- |
| Group | Brain death (h) | Age | BMI | HTN | DM | Creatinine<sup>1</sup> | Cold ischemia time (h) | Age | BMI | eGFR at 12 months |
| 1<br><10h | 5.9 +/- 4.0 | 73 +/- 8.3 | 31.7 +/- 0.5 | 2/2 HTN | 1/2 DM | 85.0 +/- 8.4 | 14.1 +/- 0.7 | 67.5 +/- 0.7 | 24.1 +/- 0.6 | 49.5 +/- 33.9 |
| 2<br>10-15h | 13.1 +/- 1.9 | 51.4 +/- 11.3 | 24.8 +/- 2.7 | 0/5 HTN | 0/5 DM | 62.4 +/- 9.8 | 13.6 +/- 5.3 | 51.6 +/- 16.5 | 22.6 +/- 5.3<sup>3</sup> | 49.2 +/- 8.5 |
| 3<br>15-20h | 17.9 +/- 1.4 | 53.6 +/- 6.2 | 31.7 +/- 6.9 | 3/5 HTN | 1/5 DM | 66.6 +/- 18.8 | 13.2 +/- 1.2<sup>2</sup> | 55.0 +/- 1.8 | 25.53 +/- 5.3<sup>3</sup> | 48.6 +/- 14.1 |
| 4<br>20-25h | 22.6 +/- 1.9 | 49.2 +/- 8.1 | 29.0 +/- 5.7 | 1/5 HTN | 1/5 DM | 59.4 +/- 24.0 | 18.3 +/- 6.0 | 47.4 +/- 19.3 | 26.4 +/- 5.0<sup>2</sup> | 65.4 +/- 44.7 |
| 5<br>25-30h | 26.5 +/- 1.6 | 48.8 +/- 10.4 | 25.9 +/- 3.4 | 1/5 HTN | 1/5 DM | 59.4 +/- 24.0 | 14.4 +/- 4.0 | 42.2 +/- 19.7 | 27.5 +/- 5.5 | 51.8 +/- 19.7 |
| 6<br>>30h | 33.0 +/- 3.6 | 36.6 +/- 15.1 | 32.7 +/- 9.8 | 1/4 HTN<sup>2</sup> | 1/5 DM | 83.8 +/- 33.0 | 18.2 +/- 6.1 | 40.- +/- 19.8<sup>2</sup> | 24.8 +/- 1.29<sup>2</sup> | 56.8 +/- 20.3 |
| P value (2-5) | <0.0001 | 0.833 | 0.16 |  |  | 0.95 | 0.70 | 0.63 | 0.74 | 0.70 |
| P value<sup>4</sup> (1-6) | <0.0001 | 0.015 | 0.28 |  |  | 0.45 | 0.42 | 0.37 | 0.72 | 0.89 |

Overview of created study cohort of n=27 donors, group 1 included 2 donors, the remaining groups included 5 donors each. The cohort was created by selecting donors with intracranial haemorrhage (ICH), a documented length of brain death and full set of serum samples; donors
were matched within each 5 hour group of brain death. Extremes of post-transplantation outcome were excluded, as were donors with documented other sources of inflammatory changes such as trauma or infection. 1 at offer 2 n=1 missing data point 3 n=2 missing data points 4 One way ANOVA; all shown as mean +/- SD.

### 3.2 Creation of time course analysis

Serum levels of IL-6, TNF-a, C5a, NSE and GFAP were recorded at time points of admission (sample DB1), after confirmation of brain death (sample DB2), and at the end of organ donor management (sample DB3) (Fig. 1A). As each sample was taken at a clinically defined time point, comparison was only possible by setting the sample taken after confirmation of brain death as t=0. The timeline was divided into the period of ICH management prior to confirmation of brain death (t<0) and a period of organ donor management (t>0) after brain death was confirmed. A direct comparison to prior studies of biomarkers of brain injury and pro-inflammatory mediators is not straightforward due to heterogeneity of underlying pathologies (traumatic brain injury, ischaemic or haemorrhagic stroke, brain injury following cardiac arrest) as well as selection of different time points in a dynamic and evolving situation across studies (see Supplementary Table^20–28^).

The described selection process of donors with similar backgrounds and identical pathologies aimed to reduce heterogeneity to create a study cohort. In addition, prospective rules were agreed to identify outliers which were then excluded before undertaking a time course analysis (see Methods). This resulted in the exclusion of three donors with outlying results for at least 4 of the 5 studied biomarkers. Two of the excluded donors were in the (additional) “long BD” group. Individual donor trajectories for the remaining 24 donors broken down by IL-6, TNF-a, C5a, NSE and GFAP are shown in Figure 1B.

Each donor has a unique duration of brain death before organ procurement, but combining the information from all 24 donor samples into a ‘moving median’ allowed us to ascertain an underlying time course. Figure 1C demonstrates the moving median with a window size of 20 hours for each of the serum markers from 20 hours prior to t=0 to 40 hours after confirmation of brain death. The moving median delineates a time course for each measured serum marker over time. The pro-inflammatory cytokines IL-6 and TNF-a do *not* follow a time course that would indicate a progressively hostile environment; TNF-a levels in serum decline from admission to end of donor management and IL-6 demonstrates a broad peak around confirmation of brain death, with subsequently decline. NSE (released from damaged neurons) and the complement cascade component C5a both remain elevated from patient admission through confirmation of brain death to end of donor management. GFAP, indicating glial cell breakdown, demonstrates a step-like time course reaching plateau over the donor management period. Donor brain death is believed to occur as a result of irreversible damage to the brain stem secondary to oedema and ultimately brain stem herniation^29^; Our findings suggest that whilst neuronal damage leads to a steady level of released NSE from admission onwards, there is a step change in glial breakdown products (detected in serum) around the time of confirmation of brain death reflective of larger volume glial injury.

## 4. Discussion

Organs from brain dead donors underperform in the long term and only function at a level comparable to organs obtained from donors after circulatory death^1^; DCD kidneys, for example, experience a higher rate of delayed graft function (DGF), however – if DGF remains brief - this does not translate into worse long-term recipient or graft survival^30^. Following confirmation of brain death whilst patients remain in critical care units, there is a logistical lag until organ procurement, we are therefore offered a window of opportunity to optimise donors further and improve organ function or reduce damage. In 2017/18 in the UK, there were on average 21 hours 48 minutes between the discussion with the donor’s family and start of the procurement ^31^. In the USA, longer donor management times are reported, which is likely a reflection of different geographical and logistical solutions to the procurement and transportation of donor organs^32^. Our study offers a first characterisation of the serum changes during the duration of donor management in UK critical care units, reporting alterations in key pro-inflammatory mediators as well as markers of central nervous system injury that are not usually present in human serum or plasma in the absence of injury^33^. The time courses of the different biomarkers obtained by combining individual donor trajectories challenge the current consensus of a progressively pro-inflammatory environment, -- the so-called cytokine storm, -- after brain death.

Our work has several strengths such as the use of high-quality clinical samples from a UK biobank with a high consent rate (>85% of all DBD donors), thus a very good representation of the overall organ donor population. Careful matching of donors, and exclusion of those where other factors could affect inflammatory markers (such as due to trauma, anoxia or infection), allowed us to infer the time-course in the ‘most typical DBD donor’: an isolated intracranial haemorrhage as cause for brain death. The samples that form QUOD biobank are collected based on clinical events, such as confirmation of brain death after brain stem testing, rather than set time points; this is of benefit in a situation where we do not understand the underlying time course.

The study has also limitations. Firstly, the final study cohort that allowed for matched characteristics was small (24 donors). This was due to the limited availability of a admission (DB1) sample, only present in around 10% of all QUOD DBD samples. Our experimental design excluded patients where pathology preceding brain death would have been likely to produce systemic or neuroinflammation (e.g. traumatic brain injury) or are likely to differ in pathology (e.g. hypoxia), as these could confound the results. It is likely that brain-death related molecular changes follow different trajectories when brain death occurs as a consequence of global hypoxia rather than intracranial haemorrhage. Finally, it should be noted that our goal was to understand changes representing serum levels of pro-inflammatory markers present in the donor and thus impacting the transplants- to-be, rather than detailed kinetic analysis including release or breakdown of molecules. Additionally the study was not designed to compare individual organ outcomes and did not include a comparison of groups with known ‘good’ vs. ‘bad’ post-transplantation outcomes, such as e.g. eGFR after 12 months.

This is the first time-course analysis of serum TNF-alpha, IL-6 and C5a in human DBD donors surrounding the donor care up to organ procurement. Serum changes of NSE and GFAP provide insights into the intracranial pathology with increasing release of proteins indicating progressive glial damage around confirmation of brain death.

Ultimately, however, the next steps are to build on our findings to determine how to improve organ quality and post-transplant outcomes. Legally and ethically, treatments to any donor aiming to improve transplantation outcomes can only be administered *after* brain death is confirmed. Therefore, understanding which pathways are amenable to intervention during this time frame is paramount. Targeting pathways or molecules that are already low or declining - whether due to the treatments provided in critical care or as part of a response to the initial injury - would be futile and likely not translate into meaningful results. Our work suggests that IL-6 or complement should be considered feasible targets for intervention during donor management. Recent animal work confirms a plateau of serum Il-6 and decline of TNF-alpha after intracranial hypertension is experimentally induced.^34^ Apart from inhibiting pro-inflammatory mediators, induction of anti-inflammatory pathways might offer alternative avenues: Histological studies of donated DBD kidneys report expression of protective, anti-inflammatory heat shock proteins (Hsp70, HO-1) alongside known pro-inflammatory mediators – with anti-inflammatory upregulation translating into protecting or restoring renal function^35^. Both strategies could lead to reduction or elimination of long-term fibrotic changes known to be linked to e.g. macrophage polarisation and complement mediated renal inflammation.^5,18^

Importantly, our study did not provide evidence which supports the previously upheld cytokine storm theory that underpins the perceived need to procure organs as quickly as possible. A recent UNOS study of cardiac transplantation showed that longer (>42 hours) brain death times did not result in worse outcomes^36^. A study from Israel found no correlation between duration of brain death and adverse outcomes in cardiac transplantation, although short duration of brain death was defined as shorter than 97 hours^37^. Our results have possible implications for further work to identify optimal procurement time points.

Finally, we believe our work translates more widely by proposing a novel approach how human samples collected e.g. as part of a biobank can be used to study time courses directly in humans and guide identification of treatment targets and timings. By creating a ‘meta-cohort’ from post hoc data, our modelling approach allowed us to use limited clinical samples to study the time-course of various biomarkers leading up to organ donation. Development of new therapies typically relies upon pre-clinical animal models. However, in cases where the underlying biology is poorly understood, human pathophysiology differs or no appropriate models exist, alternative methods are required. Our approach allows to study serum changes surrounding key events using human samples and can be translated to other clinical settings where repeat sampling is difficult, not possible or unethical.

## Data Availability

All data produced in the present work are contained in the manuscript, clarification or further information is available upon reasonable request to the authors

## List of abbreviations

BMI: Body Mass Index
C5a: Complement component 5a
DBD: Donation after brain death
DCD: Donation after circulatory death
DGF: Delayed graft function
DM: Diabetes Mellitus
eGFR: Estimated Glomerular Filtration Rate
GFAP: Glial Fibrillary Acidic Protein
Hsp70: Heat Shock Protein 70
HO-1: Heme Oxygenase 1
ICH: Intracranial haemorrhage
IL-6: Interleukin 6
NSE: Neuron Specific Enolase
QUOD: Quality in Organ Donation
TNF-alpha: Tumour Necrosis Factor alpha

## Acknowledgments

KDB was supported by the National Institute for Health Research (NIHR) with an NIHR Academic Clinical Fellowship. KDB received a pump priming research grant from the Oxford Transplant Foundation to support this project (R56956/AA001)

The authors would like to thank Dr Sergei Maslau for his help with the initial selection of the study cohort and Dr Michael Craig for his thoughts and comments on earlier versions of the manuscript. We would like to thank all organ donors and their families – none of this work would be possible without them. This study was made possible by obtaining samples and data from the UK QUOD Biobank which is a partnership programme between UK academic transplant centres and NHS Blood and Transplant.

